# Effects of Transcranial Direct Current Stimulation in Children and Young People with Psychiatric Disorders: A Systematic Review

**DOI:** 10.1101/2022.03.17.22272541

**Authors:** Lucy Gallop, Samuel J. Westwood, Yael D. Lewis, Iain C. Campbell, Ulrike Schmidt

## Abstract

**Background:** Transcranial Direct Current Stimulation (tDCS) has demonstrated benefits in adults with various psychiatric disorders, but its clinical utility in children and young people (CYP) remains unclear.

**Objective:** This PRISMA systematic review used published and ongoing studies to examine the effects of tDCS on disorder-specific symptoms, mood and neurocognition in CYP with psychiatric disorders.

**Methods:** We searched Medline via PubMed, Embase, PsychINFO via OVID, and Clinicaltrials.gov up to January 2022. Eligible studies involved multiple session (i.e. treatment) tDCS in CYP (≤ 25 years-old) with psychiatric disorders. Two independent raters assessed the eligibility of studies and extracted data using a custom-built form.

**Results:** Of 28 eligible studies (participant N= 379), the majority (*n* = 23) reported an improvement in at least one outcome measure of disorder-specific symptoms. Few studies (*n* = 9) examined tDCS effects on mood and/or neurocognition, but findings were mainly positive. Overall, tDCS was well-tolerated with minimal side-effects. Of 11 eligible ongoing studies, many are sham-controlled RCTs (*n* = 9) with better blinding techniques and a larger estimated participant enrolment (*M* = 74.7; range: 11-172) than published studies.

**Conclusions:** Findings provide encouraging evidence of tDCS-related improvement in disorder-specific symptoms, but evidence remains limited, especially in terms of mood and neurocognitive outcomes. Ongoing studies appear to be of improved methodological quality; however, future studies should broaden outcome measures to more comprehensively assess the effects of tDCS and develop dosage guidance (i.e. treatment regimens).

## 1. Introduction

It is estimated that 10-20% of children and adolescents experience mental health disorders worldwide [1], with roughly 75% of all psychiatric disorders having an onset in childhood, adolescence, or early adulthood (mid-20s) [2,3]. This period of onset coincides with sensitive periods of experience-dependent changes in brain structure and function, with evidence showing common and disorder-specific functional disorganisation of neurocognitive and affective networks in children and young people (CYP) with psychiatric disorders (e.g. [4]). This highlights the need for early detection and intervention [5], however, pharmacotherapy in CYP remains contentious across many mental health conditions [6,7] and the efficacy of psychotherapy in routine clinical settings is considerably lower than in research trials [8].

Neuromodulation techniques, particularly non-invasive brain stimulation, are a safe and promising alternative and/or adjunct novel therapy in psychiatry [9]. One particular technique, Transcranial Direct Current Stimulation (tDCS), involves the application of a constant weak direct current via electrodes placed on the scalp [10] to induce polarity-dependent changes in cortical excitability – i.e. anodal tDCS increases cortical excitability while cathodal tDCS decreases excitability [11]. tDCS is thought to shift resting transmembrane potential [12], with longer-term changes in cortical activity underpinned by N-methyl-D-aspartate (NMDA) dependent mechanisms similar to long-term synaptic potentiation and depression [13,14]. tDCS is also portable, low cost, easy to use, and has very a good tolerability and side-effect profile [15,16], all of which make tDCS ideal for use in CYP.

In psychiatry research, tDCS has been mainly administered to adults. The data have resulted in meta-analytic evidence and promising reviews of beneficial effects (e.g. [17,18]). Studies involving CYP are growing rapidly, with >20 studies published or registered as trials on clinicaltrials.gov in the past 2-years (see also, [19,20]). Despite this, attempts to consolidate findings in CYP are outdated or limited to non-systematic, narrative reviews [16,21] or systematic reviews that have focused on literature for a specific disorder, particularly neurodevelopmental disorders [22,23], or the safety and tolerability of tDCS [15]. None have examined tDCS-effects on mood or cognition in CYP with psychiatric disorders. Therefore, we followed a rigorous methodology for systematic reviews focusing on published and unpublished studies in CYP with psychiatric disorders.

Following PRISMA 2020 (Preferred Reporting Items for Systematic Reviews and Meta-Analyses) guidelines, we systematically reviewed studies investigating tDCS effects across psychiatric disorders in CYP in order to (1) evaluate the effects of tDCS on disorder-specific symptoms and impairments, (2) determine the effects of tDCS on mood and neurocognitive outcomes, (3) outline the populations and methodologies used in ongoing trials and unpublished data.

## Methods

### Protocol and registration

This study was pre-registered (see PROSPERO, ID: CRD42019158957; and [24]) and is reported in line with PRISMA 2020 guidelines [25].

### Literature search

MEDLINE, EMBASE and PsycINFO databases were searched using the following search terms: (transcranial direct current stimulation or tDCS) AND (young people, child, adolescent, young adult, youth, boy, girl, paediatric, young people and young persons) AND (neuropsychiatric disorders, autism, ADHD, schizophrenia, mood disorder, bipolar, depression, anxiety, panic, OCD, Tourette’s, PTSD, acute stress disorder, substance abuse, eating disorders, personality disorder). The search was conducted on 28/01/21 and updated on 26/01/22. The reference lists of included studies were manually searched for additional relevant studies not identified by the database search. To identify ongoing/unpublished trials, we searched World Health Organisation International Clinical Trials Registry Platform (ICTRP) registry, the National Institute of Health (NIH) registry, the European Union Clinical Trials Register, and the International Standard Randomised Controlled Trials Number (ISRCTN) registry.

### Eligibility criteria

We included all types of full-text publications written in English that reported multiple (>1) sessions of tDCS in individuals under 26-years-old at enrolment with a psychiatric disorder. We included all types of reports, studies and multi-session tDCS protocols unless the aim was basic research, protocol development, or to investigate the mechanism of action of tDCS.

### Data extraction and analysis

Two authors (LG and YL) independently screened identified records against the eligibility criteria, extracted data, and performed the quality assessment. Data extraction was performed with a custom-made form adapted from the Cochrane data collection for intervention reviews [26] (see Supplementary Material S1 for details). A meta-analysis was not feasible due to significant heterogeneity in study designs, outcome measures, and tDCS protocols.

### Quality Assessment

LG and YL independently assessed risk of bias using the Cochrane risk of bias 2.0 tool (RoB 2.0) in randomised controlled trials (RCTs) [27], and the Cochrane tool for risk of bias in non-randomised studies of interventions (ROBINS-I) [28]. Inter-rater agreement was 90%. Conflicts were resolved by discussion.

## Results

We identified 28 eligible studies (total N= 379; age range 2-25 years, M=14.94, SD= 4.24; 82.3% male; see Fig. 1), composed of six double-blind RCTs, one double-blind, sham-controlled trial, two single-blind RCTs, one single-blind controlled clinical trial, four open-label studies, and 14 case series/studies (see Table 1 and 2). Six studies were in ASD [29-34], five in ADHD [35-39], four in schizophrenia [40-43], two each in OCD [44,45], Tourette’s syndrome [46,47], depression [48,49], anxiety [50,51], or substance abuse disorder [52,53], and one each in eating disorders [54], catatonia [55], or co-comorbid ASD, ADHD and anxiety [56]. Across studies, tDCS was typically delivered for 20 to 30-mins (M=23; SD = 4.43) once-daily over 4 to 28 sessions (M=12; SD = 7.07). However, several studies applied tDCS in twice-daily sessions [40,43-46] and one case report suggests that tDCS is an ongoing treatment [40]. Montage configurations included anodal *(n* = 14), cathodal *(n* = 2), and bilateral (*n* = 12) tDCS, with anodal tDCS to the left-DLPFC being the most frequently employed montage across studies (*n* = 8). tDCS was delivered at a stimulation intensity between 0.25mA-3mA, with 1mA (*n* = 11) or 2mA (*n* = 11) most employed.

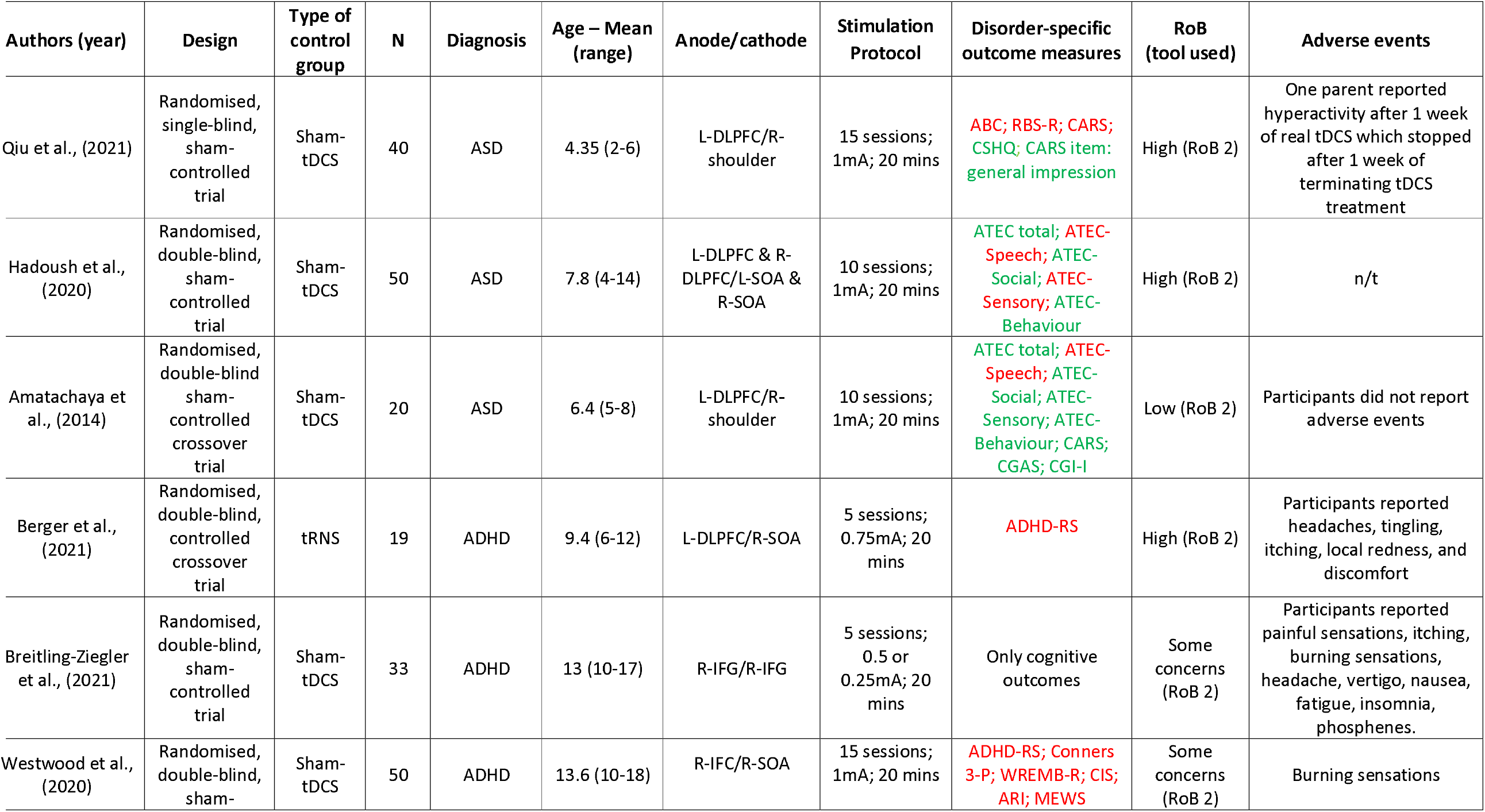
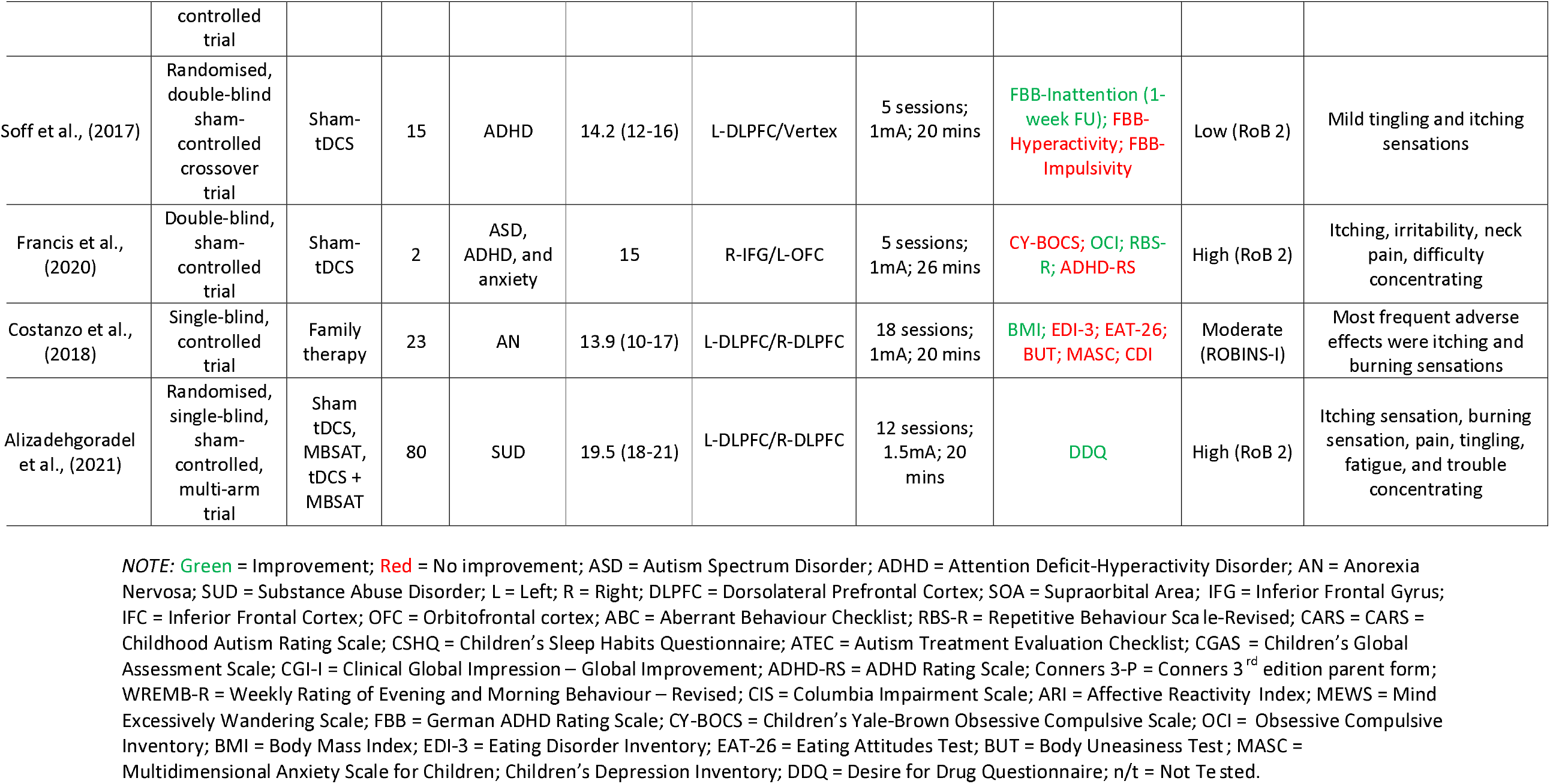

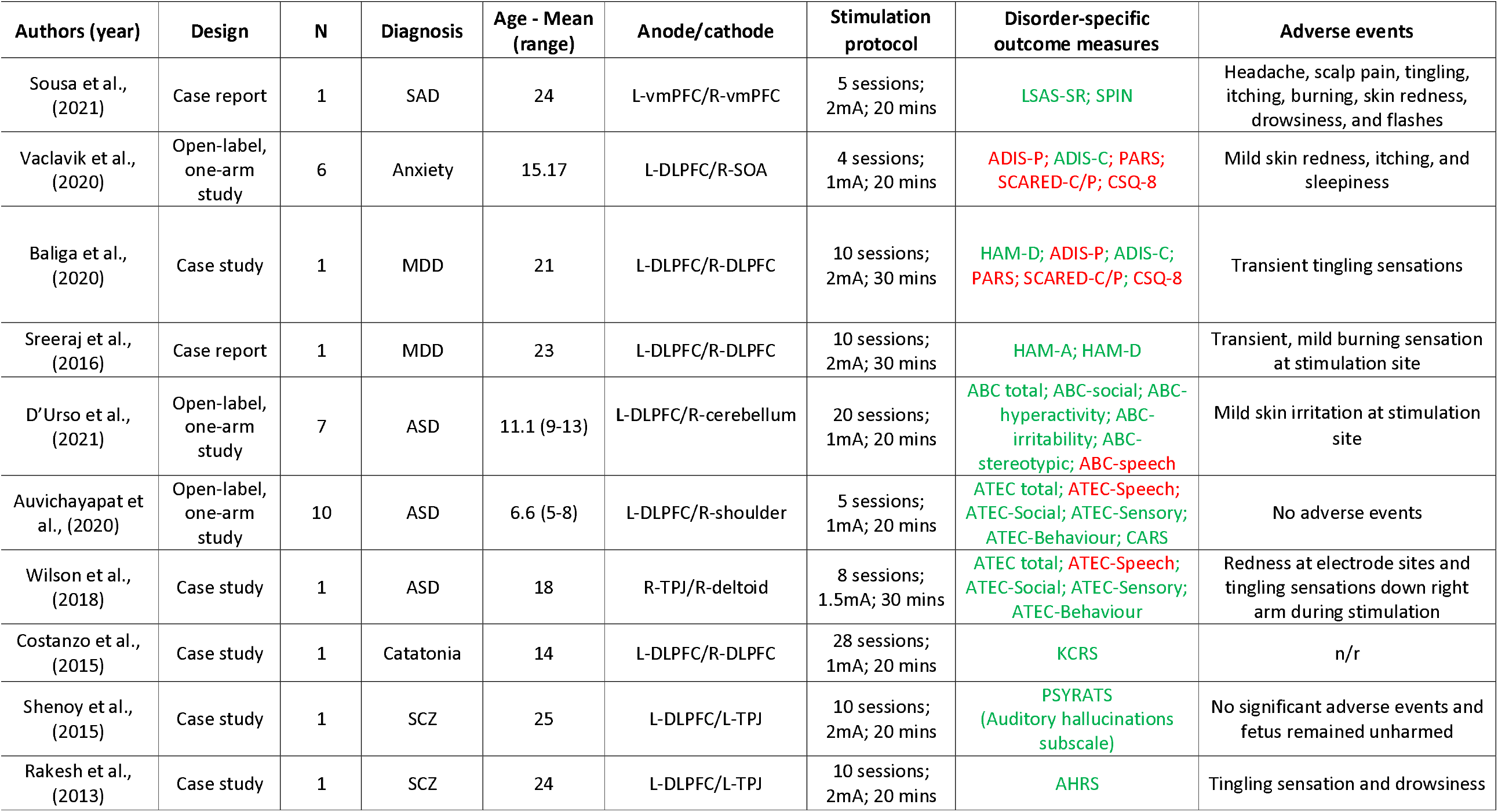
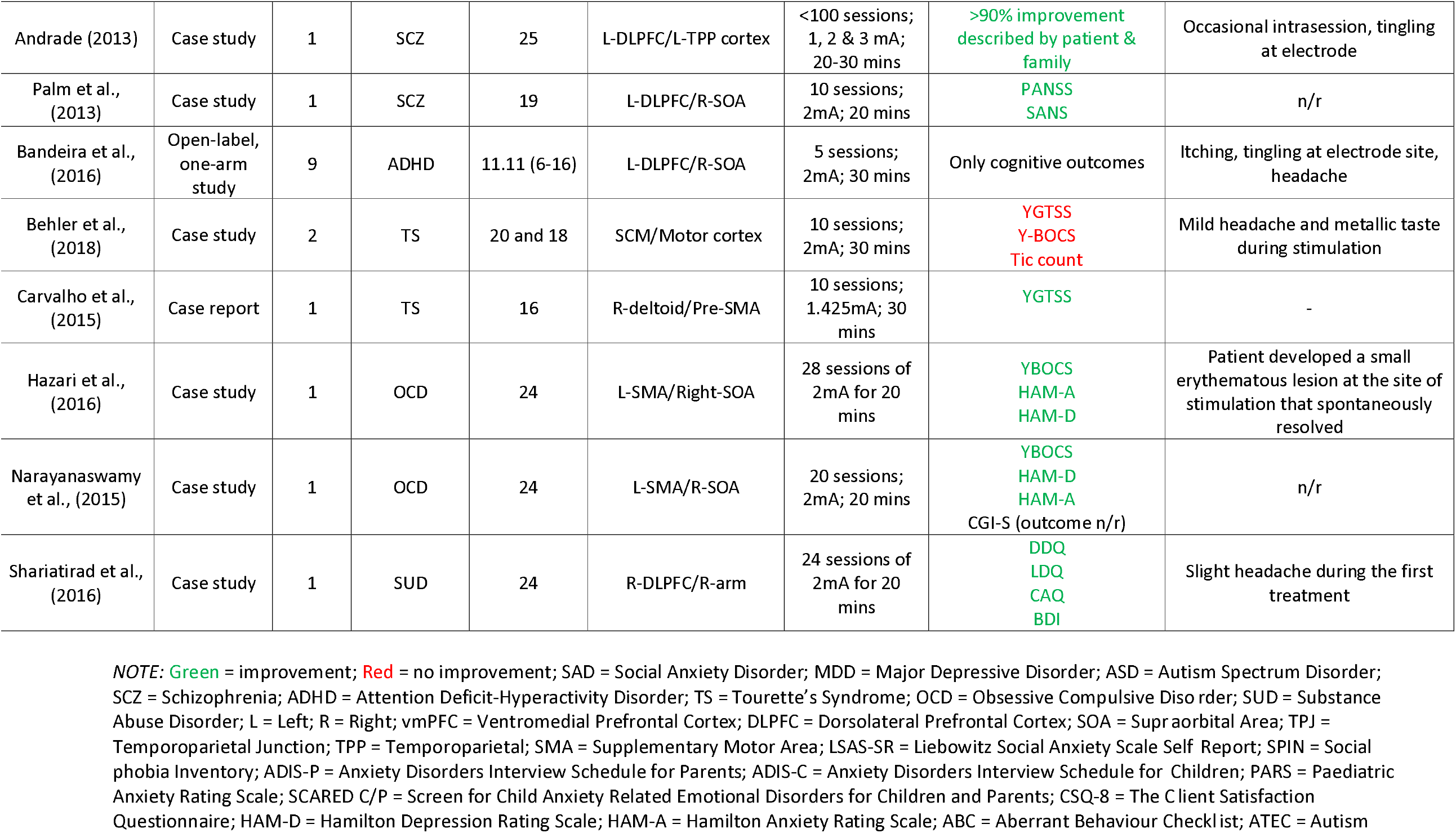
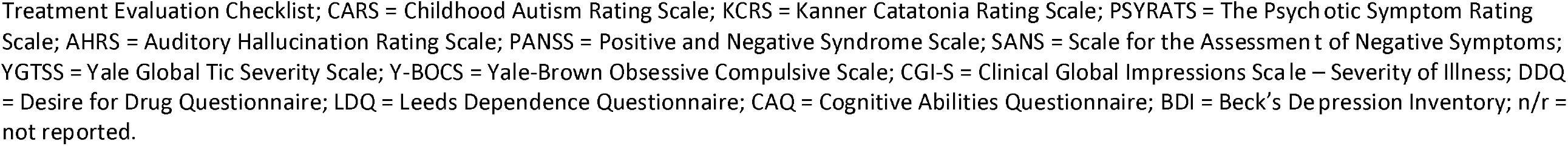

**Figure 1.**
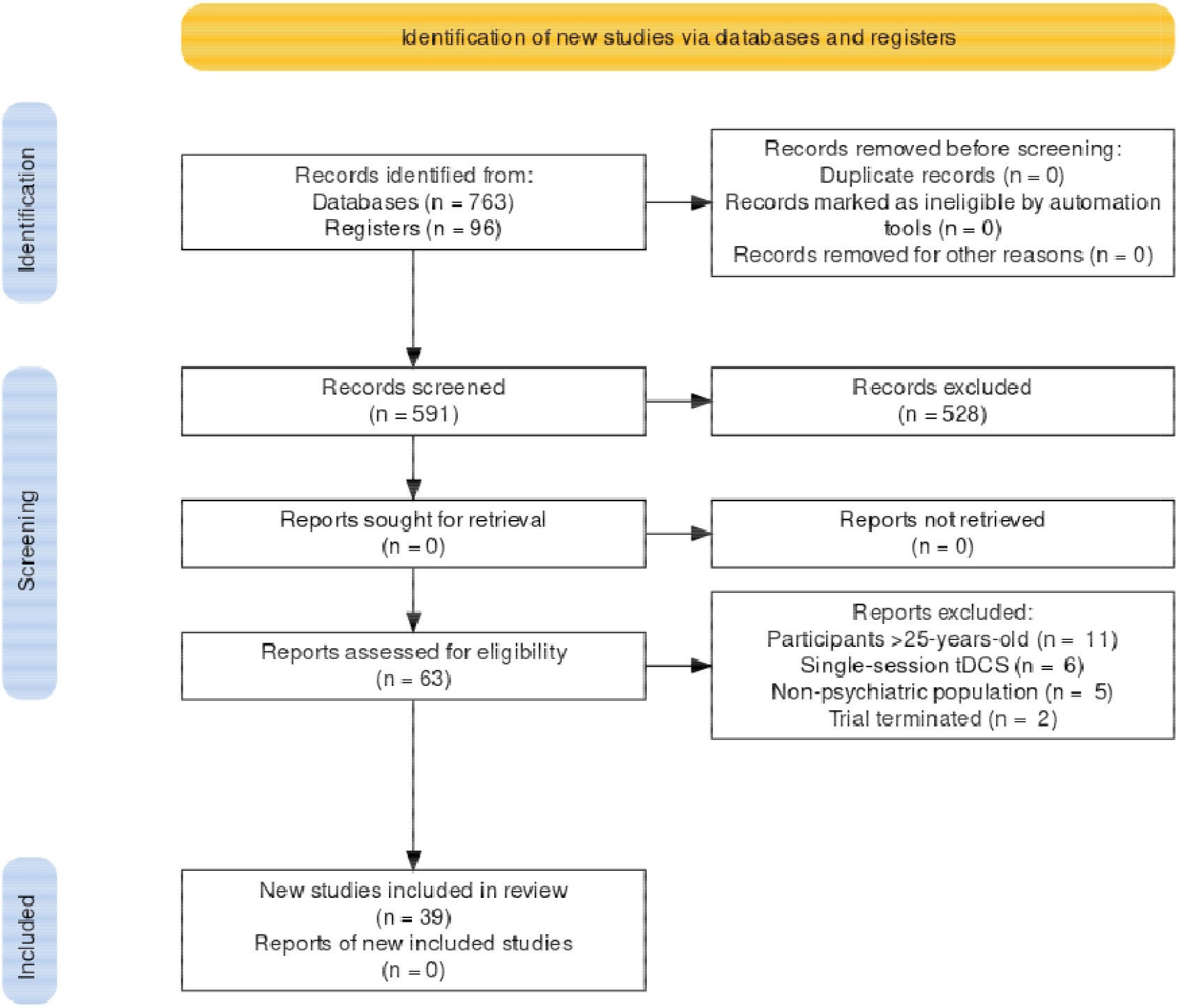
PRISMA flow diagram of selected studies (n = number of articles). A total of 28 studies were systematically reviewed and 11 ongoing/unpublished trials were identified.

### Quality Assessment

Of the nine RCTs, overall risk-of-bias was rated as “high” in five studies [32,33,36,52,56]; two with “some concerns” [37,39], and two with “low” [29,38] (see Supplementary Material S2). The non-randomised, controlled clinical trial was rated with moderate risk of bias [54]. All open-label studies, case series, and case reports were rated as low quality. Three of the nine RCTs were prospectively registered and one was retrospectively registered. Comparison of the registered protocol and final publication showed that two studies omitted a registered primary outcome ([52]; dot-probe task and difficulties in emotion regulation scale); or secondary outcomes ([36]; Behaviour Rating Inventory of Executive Function (BRIEF) and only partial reporting of Wechsler Intelligence Scale for Children (WISC-IV), and one promoted one outcome registered as “other” to a secondary outcome in the final publication ([36]; Clinical Global Impression – Severity).

### 1. Clinical tDCS Effects in Psychiatric Disorders

#### Neurodevelopmental Disorders

##### (a) Autism Spectrum Disorder (ASD)

A recent sham-controlled RCT in 43 children with ASD [32] applied 10-sessions of bifrontal anodal tDCS to the left-and right-DLPFC and reported a significant reduction in therapist-rated total ASD and related symptoms, including sociability, physical health and behaviour, but not sensory and cognitive awareness or language, compared to sham. Another recent single-blind RCT [33] in 40 children with ASD reported a significant improvement in sleep and observer-rated overall ASD severity immediately after 15-sessions of anodal tDCS over the left-DLPFC compared to sham, while ASD symptoms and related behaviour impairments remained unchanged [33].

Two studies applied 5-days of anodal tDCS over the left-DLPFC: one crossover RCT in 20 children with ASD [29] reported a significant improvement in investigator-and parent-ratings of ASD symptoms, as well as parent-rated sociability, physical health and behaviour, sensory and cognitive awareness compared to sham, but not speech and language communication, at 1-week post-stimulation. Ratings of psychosocial functioning also improved at 1-week post-stimulation compared to sham, while clinical impression of improvement was rated as “much improved” in nine and “minimally improved” in eight children [29]. A single-arm open-label study [30] in 10 children with ASD reported a significant reduction in investigator-rated ASD symptoms immediately, 1-week, and 2-weeks post-treatment, compared to baseline, whereas parent-ratings of ASD symptoms relating to sociability, physical health and behaviour, sensory and cognitive awareness, but not speech and language communication, were significantly reduced at post-treatment only [30].

A single-arm open-label study [31] applied 20-sessions of cathodal tDCS over the right cerebellar lobe in seven children with ASD, which reduced caregiver-rated aberrant behaviour symptoms one-week after stimulation compared to baseline in all but two participants, both of whom were taking psychotropic medication during the study period. Unexpectedly, one week after stimulation, one patient with a history of epilepsy showed no EEG-related epileptic activity in the frontal region, while another participant with comorbid tic disorder showed fewer, less intense tics, which remained until 3-month follow-up [31].

Lastly, a case report in an 18-year-old male with ASD applied 8 sessions of anodal tDCS to the right temporoparietal junction and reported fewer total ASD symptoms immediately and 2-months after stimulation compared to baseline [34].

##### (b) Attention Deficit Hyperactivity Disorder (ADHD)

Two double-blind RCTs applied tDCS to the inferior frontal cortex (IFC). The largest RCT in CYP with ADHD *(n* = 50) administered 15 sessions of anodal or sham tDCS to the right-IFC with concurrent cognitive training [39]. Compared to sham, the tDCS group showed significantly higher parent-rated ADHD symptoms immediately after stimulation, but not at a 6-months follow-up. No significant differences were reported in other measures of ADHD symptoms or related impairments [39]. The other RCT [37] administered five sessions of bilateral high-definition tDCS to the right-and left-IFC in 33 children and adolescents with ADHD, with stimulation intensity titrated post-randomisation to .25mA (*n* = 11) and .5mA (*n* = 9) to minimise discomfort. Results showed a significant reduction in self-rated ADHD total and hyperactivity symptoms compared to sham at post-treatment, but not at follow-up, while self-rated inattention or impulsivity and all parent-rated ADHD symptoms remained unchanged [37].

A smaller double-blind, crossover RCT [38] in 15 adolescents with ADHD applied five sessions of anodal over the left-DLPFC. Compared to sham, parent-rated inattention improved at 1-week follow-up, but not immediately after anodal tDCS. No other clinical effects were found [38]. Another double-blind, crossover RCT [36] in 19 children with ADHD compared five sessions of transcranial random noise stimulation (tRNS) over the left-DLPFC and right-IFC with tDCS to the left-DLPFC, both with concurrent executive function training. Findings showed that relative to tDCS, tRNS significantly reduced parent-rated ADHD symptoms immediately and one week after stimulation (adjusting for baseline scores), but with no difference in global clinical impressions [36].

Lastly, in a single-arm open-label study [35] in nine children with ADHD, five sessions of anodal tDCS over the left-DLPFC were combined with a picture association cognitive training task and parents reported overall improvements in behaviour. However, without a sham-control, a placebo effect cannot be ruled out.

##### (c) ASD and ADHD

A double-blind, parallel, sham-controlled case report applied 10-sessions of anodal-tDCS over the right inferior frontal gyrus (rIFG) combined with cognitive training in two 15-year-old, female, fraternal adolescent twins with ASD, ADHD, and anxiety. Both twins had parent-reported compulsive symptoms, and one had comorbid OCD. Findings showed reduced parent-rated compulsive and repetitive/restrictive symptoms, but not clinician-rated OCD symptoms or parent-rated ADHD symptoms [56].

##### (d) Tourette’s syndrome

In one case study [47] in an 18-year-old female and 20-year-old male with motor and vocal tics, bilateral cathodal tDCS was applied to the right-and left-pre-SMA twice-daily over 5 consecutive days. Compared to baseline, one participant showed a reduction in tics immediately after stimulation, whilst the other participant showed an increase in tics and OCD symptoms. Both participants self-rated negative affect decreased, and positive affect increased, but none of these changes were statistically tested [47]. One case study in a 16-year-old boy with refractory Tourette’s syndrome showed reduced motor and vocal tics 1-, 2-, 12-, and 24-weeks (relative to baseline) after 10-sessions of cathodal tDCS over the left pre-SMA [46].

#### Schizophrenia-Spectrum Disorders

##### (a) Schizophrenia

Three case studies applied bilateral tDCS (anode-left-DLPFC; cathode-right-temporo-parietal junction) over 5 consecutive days. In two of these studies, twice-daily tDCS reduced auditory hallucinations entirely at post-treatment in a 24-year-old male [42] and almost entirely 1-month after stimulation in a 25-year-old pregnant female [43], whereas, in the third study, once-daily tDCS reduced auditory hallucinations immediately after stimulation in a 25-year-old female, but she required continued once-or twice-daily stimulation to maintain improvements over three-years [40]. Lastly, one case study in a 19-year-old-male reported reduced positive and negative symptoms, disorganization, flattened affect, lack of concentration, and impetus after 10 sessions of anodal tDCS over the left-DLPFC [41].

##### (b) Catatonia

A case study in a 14-year-old female with ASD and catatonia showed reduced catatonic symptoms compared to baseline immediately and 1-month after 28-sessions of bilateral tDCS (anode-right; cathode-left) over the DLPFC [55].

#### Major Depressive Disorder

Two case studies applied 10-sessions of bilateral tDCS over the left-and right-DLPFC. One study reported that a 21-year-old male presenting with a moderate depressive episode showed fewer depressive symptoms immediately after 5-sessions of stimulation applied on two occasions separated by two-years [48]. The second reported that a 23-year-old pregnant female with a depressive episode showed fewer depressive and anxiety symptoms one-month after stimulation compared to baseline, which, the authors suggested was indicative of clinical remission [49].

#### Anxiety Disorders

A single-arm open-label study in six adolescents with social anxiety disorder or generalized anxiety disorder reported that combined anodal-tDCS to the left-DLPFC with attention bias modification training reduced self-and parent-reported total anxiety symptoms, self-and parent-rated anxiety related emotional symptoms, and clinician-rated anxiety symptoms, compared to baseline [51]. However, only self-reported total anxiety symptoms significantly reduced from baseline to post-stimulation.

A case study in a 24-year-old female with social anxiety disorder reported reduced self-reported social anxiety symptoms immediately after 5-sessions of anodal tDCS over the left vmPFC, and at 15-days follow-up, although improvements did not reach clinical significance [50].

#### Obsessive Compulsive Disorder (OCD)

Two case studies applied anodal tDCS over the left pre-SMA/SMA twice-daily over 10-conseuctive days. In one, a 24-year-old male with OCD and co-occurring depressive symptoms showed fewer OCD symptoms compared to baseline immediately and 7-months after stimulation [44]. The same stimulation protocol was applied for 8-sessions two-years later, which reduced recurring OCD and depressive symptoms, but also lesioned the skin under the stimulation site [44]. The second case study reported a reduction in OCD, depression, and anxiety symptoms at one week and 1-month after stimulation in a 24-year-old male [45].

#### Substance Abuse Disorder

A single-blind, parallel group RCT in 80 boys with methamphetamine addiction administered 12-sessions of anodal tDCS over the left-DLPFC with or without combined mindfulness training, mindfulness training only and sham tDCS only [52]. The results showed a significant reduction in desire for drugs at post-treatment and 1-month follow-up in the three active treatment groups, but not in the sham group, compared to baseline [52].

In a case report in a 24-year-old male with methamphetamine use disorder, anodal-tDCS over the right DLPFC reduced self-reported drug cravings immediately after 20-sessions of stimulation and, after four more sessions, at 6-months follow-up, at which point paranoid delusions and hallucinations had also reduced completely [53].

#### Eating Disorders

One clinically controlled trial [54] in 23 adolescents with Anorexia Nervosa (AN) combined treatment as usual with either family therapy or 18-sessions of bilateral tDCS over the DLPFC (anode-left; cathode-right). It was not clear how people were allocated to these two study arms. Compared to baseline, only the tDCS group had significantly reduced BMI immediately after stimulation and at 1-month follow-up. Both groups showed significantly reduced overall AN, depression, and anxiety symptoms compared to baseline, but no significant Group by Time interaction, and thus, placebo effects cannot be ruled out [54].

### 2. tDCS Mood Effects

#### OCD

Two case studies reported reduced clinician-rated depression symptoms on the Hamilton Depression Rating Scale at post-stimulation (relative to baseline) [44,45].

#### Substance Abuse Disorder

In a case report [53], the participant self-reported reduced depression, on the Beck Depression Inventory, at post-stimulation, 3-months and 6-months follow-up.

### 3. Neurocognitive tDCS Effects in Psychiatry Disorders

#### ADHD

A double-blind RCT [37] comparing 0.5mA and 0.25mA to sham reported a significant reduction in reaction time variability in a combined Go/No-Go and n-back task immediately and 4-months after 5-sessions of 0.5mA anodal HD-tDCS over the right IFC. In contrast, in the same task, the 0.25mA group showed an increase in no-go commission errors over the course of tDCS, and this effect became significant at day-5, but was non-significant at post-stimulation and at 4-month follow-up. At post-stimulation, the 0.5mA group also showed a significant reduction in reaction time variability in the flanker task and a reduced number of commission errors in the spanboard task compared to sham, but neither effect was significant at follow-up [37].

One double-blind, crossover trial [38] showed that compared to sham, anodal tDCS improved QbTest (a combined working memory (n-back minus-2) and go/no-go task) measures of attention at 7-day follow-up, but not immediately after stimulation, in measures of hyperactivity immediately and 7-days stimulation, but not in measures of impulsiveness. In a double-blind, crossover RCT [36], tRNS improved working memory, but not short-term memory, and only processing speed in a sustained attention task, compared to tDCS. In addition, exploratory moderation analysis predicted a trend-level larger tRNS effect in parent-rated ADHD symptoms for participants with the greatest working memory improvement.

In a double-blind, parallel RCT [39], there were no significant effects of tDCS across measures of motor and interference inhibition, time estimation, sustained attention, cognitive flexibility, visuospatial working memory, and three task-independent measures processing speed, intrasubject response variability, and prematurity. Finally, a single-arm open-label study [35] reported a significant reduction in errors on attention (omission) and switch tasks compared to baseline, but no improvement in verbal or visuospatial working memory.

#### Substance Abuse Disorder

A single-blind, parallel RCT [52] reported a significant group by time interaction in n-back task reaction times and accuracy, Wisconsin Card Sorting Task perseverative errors, and in a risk-taking task, all due to an improvement from baseline to immediately and 1-month after tDCS only, tDCS with mindfulness training, or mindfulness training only, but not sham. There was no equivalent significant effect on any of the go/no-go task measures.

In a case report [53], in a questionnaire assessing cognitive abilities, the patient showed consistent improvements from baseline at 2-months, 4-months, and 6-months post-stimulation on subscales measuring memory, inhibitory control, selective attention, decision making, planning, sustaining attention, and cognitive flexibility, but not in social cognition.

#### Schizophrenia

One case report showed that compared to baseline the patient had faster completion time on the Trail Making Test (TMT) Part A and B and fewer errors on the Self-Ordered Pointing Task (SOPT) one-and two-weeks post stimulation [41].

##### Safety

Overall, tDCS was well-tolerated and feasible in a variety of age groups and psychiatric disorders, which extends existing evidence of a good side-effect and tolerability profile of tDCS in children and adolescents [15]. However, there were exceptions, e.g. a report of lesions at the site of stimulation [44], which should be considered when developing stimulation protocols.

### 5. Unpublished Registered Trials

Of the 13 registered trials (see Table 3), one was withdrawn (unable to recruit any participants), and one was terminated (due to lack of funding). Of the remaining eleven studies (n=2 completed in August 2020 and October 2021; the rest ongoing), two are open label (one single-arm; one-RCT) and nine are double-blind RCTs trials. These 11 studies are a) recruiting either ASD (n=5), ADHD (n=4), or depression (n=2); b) stimulating the DLPFC (n=8) or temporal parietal junction (n=1; n=2 did not report stimulation site); c) applying tDCS alone (n=6), or combining stimulation with cognitive training (n=4) or applied behaviour therapy (n=1); and e) recruiting ∼70 participants (range: 11-172) per trial, in CYP aged (on average) between 10-18-years.

**Table 3.**
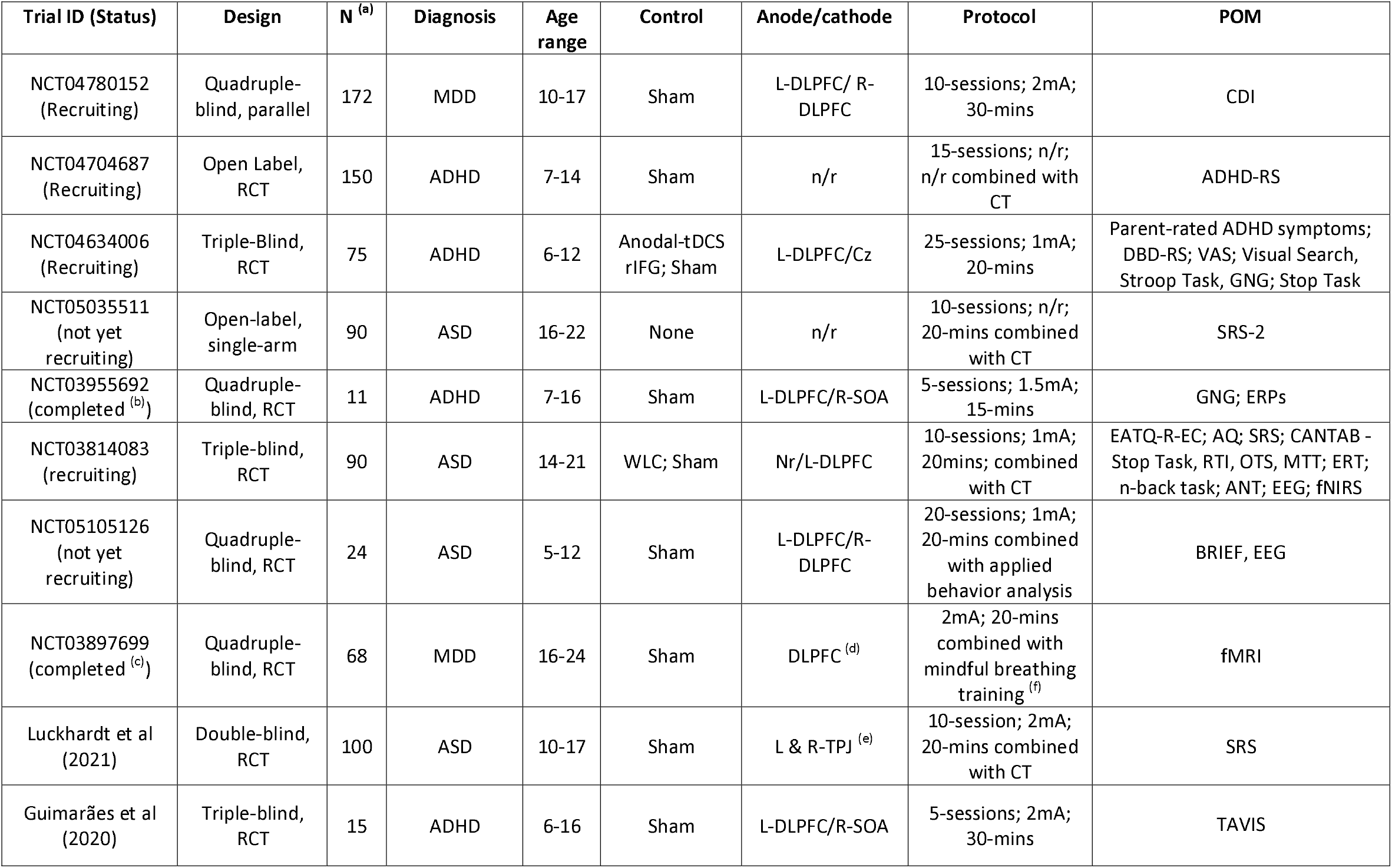
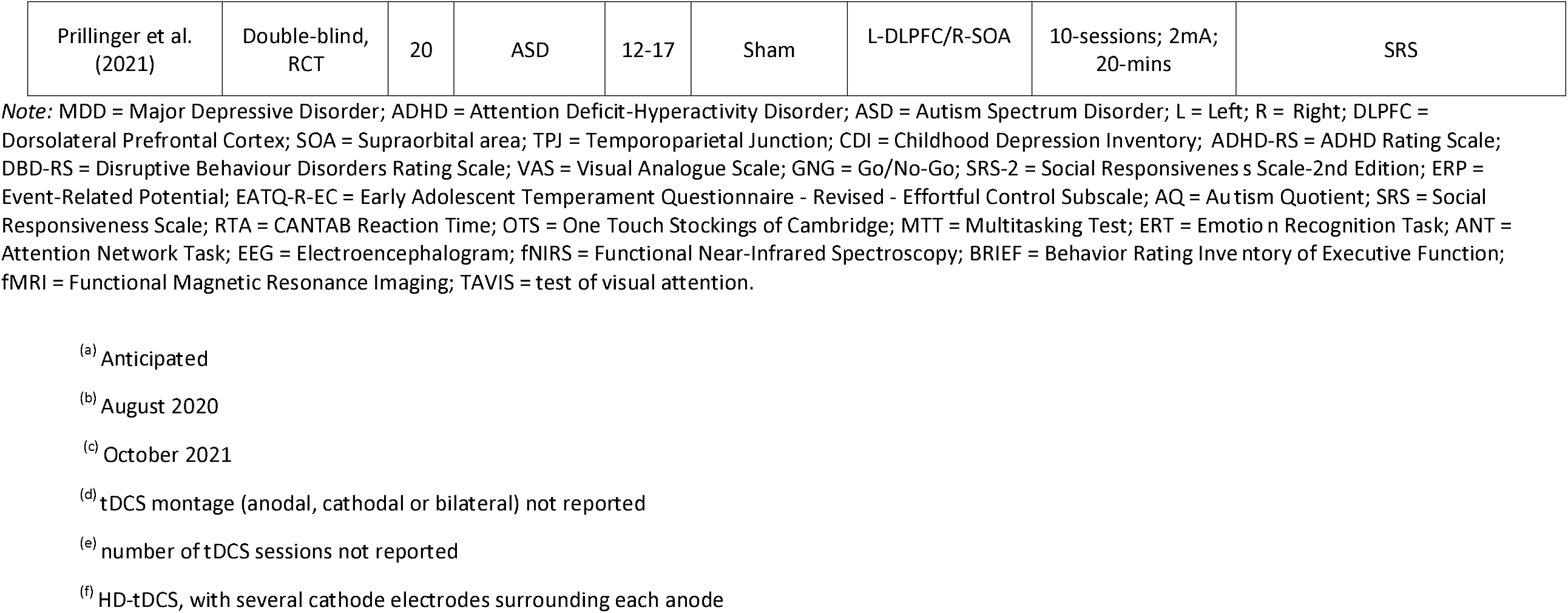
Ongoing or upcoming trials.

## Discussion

This is the first systematic review that collates published and unpublished studies investigating the effects of multi-session tDCS applied to CYP with psychiatric disorders. To date, studies are limited to case reports (n=16) and small RCTs (n=6) that demonstrate tDCS is well-tolerated, and that it is feasible to conduct RCTs in CYP, particularly those with ADHD and ASD. There is some encouraging evidence of improvement in clinical, cognitive, or mood measures, however, it is not possible to determine the therapeutic efficacy of multi-session tDCS for CYP with psychiatric disorders.

Of the 28 included studies, 26 measured clinical effects immediately after the final tDCS session, with all except three [38,39,46] reporting an improvement in at least one outcome measure of core disorder-specific symptoms. Of the ten studies that measured clinical effects at a longer-term follow-up, improvements in core symptoms persisted at 1-week [30], 2-weeks [30], 1-month [43,45,49,52,54,55], 6-months [53], or 7-months [44] after the final session of tDCS, with one study reporting no effect at 6-months [39]. Overall, these findings are in line with evidence of improvement in clinical outcomes in adults with psychiatric disorders, which have been shown to persist up to at least one-month post-stimulation (e.g. [18]). Interestingly, clinical effects persisted with a maintenance dose in one case [40] or were significantly improved at 1-week but not immediately after stimulation [38]. This might be related to evidence showing that the effects of tDCS can be delayed and only emerge after the acute treatment phase [57]. On this basis, future studies should assess the core and related symptoms of psychiatric disorders in CYP both immediately and at one or multiple longer-term follows.

Few studies measured neurocognitive or mood outcomes. Eight studies measured neurocognitive outcomes, of which four found tDCS-related improvements in the first assessment immediately [35,37,38,52], 1-week [41], or 1-month [53] after the last stimulation session, which persisted in five of the studies until 1-week [38], 2-weeks [41], 1-month [52], 4-months [37], and 6-months [53] follow-up. Two studies reported no effect of tDCS on cognition compared to sham [39] or tRNS [36] at post-stimulation and/or follow-up. All three studies measuring mood outcomes found improvements immediately after tDCS [44,45,53], with one study testing and also finding the effect at 6-months follow-up [53].

In the ten studies with cognitive and/or mood outcomes, all except three [39,44,45] stimulated the DLPFC. This is in line with substantial meta-analytic evidence that the DLPFC subserves executive functions or regulates mood [e.g. [58-61]), and with tDCS studies in healthy controls, which have reported improved cognitive outcomes up to 12-months [62] and in mood outcomes [63] post-stimulation. Twelve other studies also stimulated the DLFPC, but none measured cognitive or mood outcomes. Impaired executive functioning and mood regulation mediate the pathophysiology of many psychiatric disorders (depression [64], schizophrenia [65], ADHD [66], and ASD [67]). Further, patients often desire alternative treatments that improve executive functioning or mood over symptoms [68] without side effects associated with pharmacological interventions, such as secondary blunted affect [69], weight gain and poor social functioning [70]. It is therefore important that future research measure the effects of tDCS on a variety of disorder-relevant cognitive outcomes and mood impairments.

Heterogeneous stimulation protocols and lack of dosage-guidance meant we were not able to identify optimal stimulation parameters in CYP with psychiatric disorders. This is of concern given the neurophysiological effects of tDCS may be non-linear, and because emerging evidence suggests tDCS may modulate cortical excitability via the scalp and/or peripheral nerves, which may complicate predictions about the dose-response relationship and the reproducibility of findings [71]. For example, in one RCT [37], adolescents with ADHD received sham (n=13), or 0.5mA (n=9), or 0.25mA (n=11) anodal HD-tDCS to the IFC depending on cutaneous sensitivity. Compared to sham, the 0.25mA group showed significantly reduced response inhibition, an effect not observed in the 0.5mA group, which opposed the authors hypothesis that increasing right-IFC activity would improve executive function. In addition, in one case series [46], cathodal tDCS to the motor cortex did not improve symptoms in Tourette’s syndrome, and actually increased tic-count, whereas in another case report [47], cathodal tDCS to the pre-SMA improved both motor and verbal tics. The majority of studies (*n* = 22) used a stimulation intensity of 1-2mA, based on studies in adult populations. However, the stimulation intensity required to modulate cortical excitability in a polarity-dependent manner [72] or induce longer-term effects that persist after stimulation cessation in CYP differs from that in adults. Finally, there is evidence that of indirect stimulation of non-target sites [73], leaving the possibility of unintentional modulation of symptoms, behaviour or cognition in a potentially clinically meaningful manner. For example, D’Urso et al., [31] unexpectedly found improvements in epileptic activity and comorbid tic disorder in two participants with ASD following cathodal tDCS to the right cerebellar lobe. Overall, these inconsistent or unexpected findings (i.e. [31,37,46,47]) underline the need to improve understand the underlying biophysiological mechanisms of tDCS, as well as how different parameters (e.g. stimulation intensity) interact with the tissue being stimulated. One way to address this and identify optimal parameters is for future studies to broaden outcome measures to capture potential unintended effects on regions functionally related to target areas, and to explore the parameter space ideally using Bayesian optimisation (e.g. [74]), focal forms of tDCS (e.g., HD-TDCS), or open-source computational modelling software (e.g., ROAST; [75]).

## Limitations

Interpretation is constrained by methodological limitations present in included studies. Heterogeneity in study design, outcome measures, stimulation protocols, and participant characteristics (e.g., age, gender, disorder profile) limited comparisons across findings and adequately powered meta-analyses of clinical, cognitive, or mood outcomes. All open-label studies and case reports or series were rated as poor quality, while RCTs had some concerns (n=2) or a high (n=5) risk of bias. These ratings were mainly due to a lack of detail regarding randomisation and allocation concealment. Only 13 studies performed any statistical analysis on outcome measures, and of those studies that reported statistically significant effects, four were open-label or case series/reports. Across studies, one [39] corrected for multiple testing and assessed integrity of blinding of raters and/or experimenters, i.e. it cannot be ruled out that effects were due to placebo or test-retest effects, false-positives, and/or bias by knowledge of group assignment [76]. Only five studies combined stimulation with cognitive training, which has been reported to boost and prolong the effects of tDCS [77,78]. Sample sizes were between 15 and 50, which is short of that required to detect a medium effect in cognitive tasks (e.g. [79]).

It appears that the direction of traffic is towards improving the quality of studies. This is reflected by the fact that the majority of the 11 ongoing, or upcoming, trials we identified are double-, triple-, or quadruple-blind RCTs with larger sample sizes (∼70 on average) with nearly half combining tDCS with cognitive training across multiple sessions. However, 9 out of 11 registered trials are recruiting children with either ASD or ADHD, thus we cannot be sure that the same improvement in study quality will be seen across other psychiatric disorders or non-registered trials.

## Conclusion

Although encouraging, the evidence to date is insufficient to conclude that tDCS can improve clinical symptoms, mood, or cognition in CYP with psychiatric disorders. This is largely due to the heterogeneous study designs, limited outcomes, and small sample sizes, as these limit the interpretability and comparability of findings across studies. Future studies should seek to confirm existing findings with larger samples, and randomised, sham-controlled designs that include measures of clinical, cognitive, and mood outcomes immediately after stimulation and in longer-term follow-ups. Stimulation protocols should be justified and should consider any possible unintended outcomes that might occur, particularly in younger populations.

## Data Availability

All data produced in the present work are contained in the manuscript

## Acknowledgements

LG is supported by a PhD studentship from the National Institute of Health Research (NIHR) Mental Health Biomedical Research Centre (BRC) at South London and Maudsley NHS Foundation Trust (SLaM) and King’s College London (KCL). YDL is supported by the Daniel Turnberg Royal College of Physicians travel fellowship. US receives salary support from the NIHR BRC for Mental Health, SLaM, and Institute of Psychiatry, Psychology and Neuroscience, KCL and is also supported by an NIHR Senior Investigator Award. The views expressed in this publication are those of the authors and not necessarily those of the National Health Service, the NIHR, or the UK Department of Health.

