## Supplementary Material for "Effects of Transcranial Direct Current Stimulation in Children and Young People with Psychiatric Disorders: A Systematic Review"

*Supplementary Material S1: Data extraction*

We extracted the following information: (a) study characteristics (e.g. article title, reference, study design, sample size calculation); (b) participants (e.g. sample size, age, gender, inclusion/exclusion criteria, main disorder, illness severity, comorbidities); (c) tDCS (type of tDCS used (i.e. anodal, cathodal), site of stimulation, stimulation intensity, duration, total number of sessions); (d) comparators (sham, treatment as usual, waitlist, no comparison); (e) concurrent treatment (medication, psychotherapy, cognitive remediation); (f) outcomes (disorder-specific symptoms, mood, cognition, adverse effects).

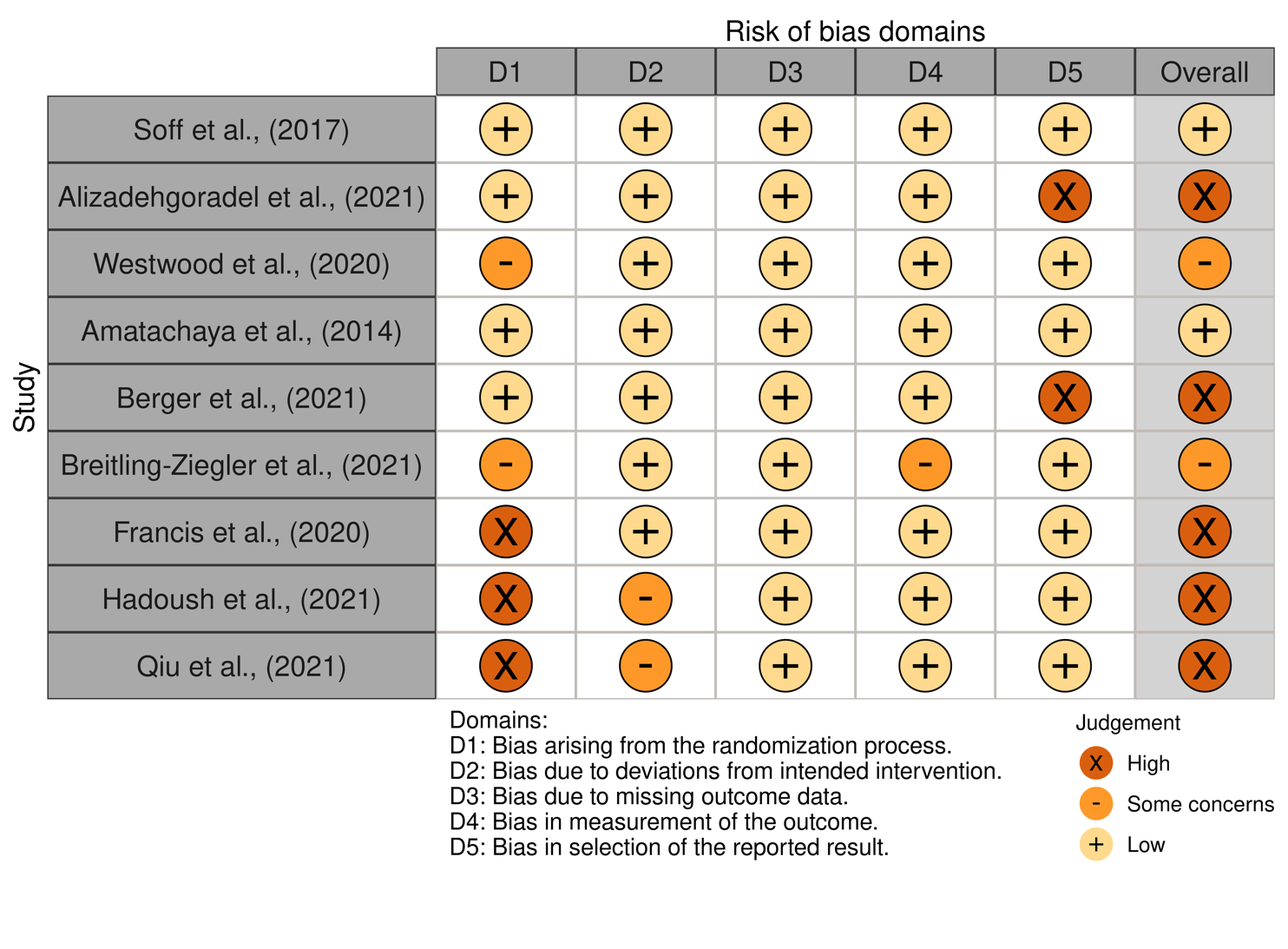
*Supplementary Material S2: Results of risk of bias assessment*

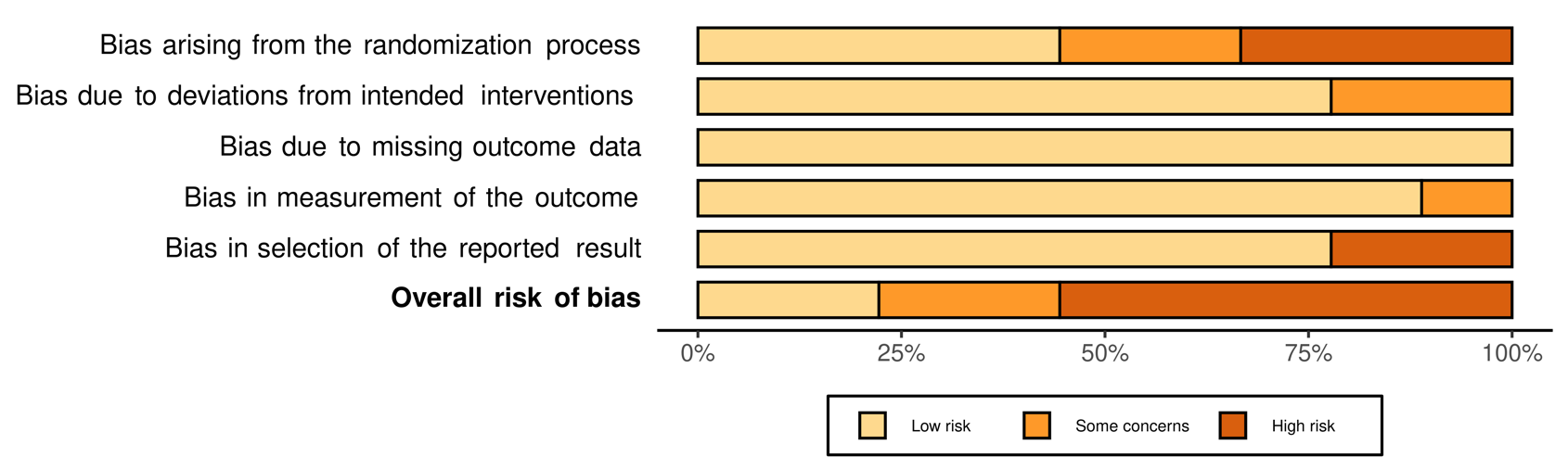
